# Effect of nutrition assessment, counselling and support integration on mother- infant nutritional status, practices and health in Tororo and Butaleja districts, Uganda: A comparative non equivalent quasi experimental study

**DOI:** 10.1101/2023.12.20.23300314

**Authors:** Samalie Namukose, Gakenia Wamuyu Maina, Suzanne N Kiwanuka, Fredrick Edward Makumbi

## Abstract

**Background:** Malnutrition remains a health challenge for women aged 15 to 49 years and their infants. While Nutrition Assessment Counselling and Support (NACS) is considered a promising strategy, evidence on its effectiveness remains scanty. This study assessed the effect of comprehensive NACS package on the mother-infant practices, health and nutrition outcomes in two districts in Eastern Uganda.

**Methods:** A comparative non equivalent quasi experimental design was employed with two groups; Comprehensive NACS (Tororo) and Routine NACS (Butaleja). Pregnant mothers were enrolled spanning various trimesters and followed through the antenatal periods and post-delivery for health and nutrition status. Infants were followed for feeding practices, health and nutritional status at birth and weeks 6, 10, 14 and at month 6, 9 and 12 post-delivery. Propensity score matching ensured study group comparability. The NACS effect was estimated by nearest neighbour matching and the logistic regression methods. Statistical analysis utilised STATA version 15 and R version 4.1.1.

**Results:** A total of 666/784 (85%) with complete data and were analysed (routine: 412, comprehensive: 254). Both groups were comparable by mothers’ age, MUAC, prior antenatal visits, meal frequency, micronutrient supplementation and instances of maternal headache, depression and diarrhoea. However, differences existed in gestation age, income, family size, education and other living conditions. Comprehensive NACS infants exhibited higher infant birth weights, weight- for- age z-scores at the 3^rd^ -6^th^ visits (p<0.001), length- for- age z scores at the 4^th^ -7^th^ visits (p<0.001) and weight-for-length z-scores at the 3^rd^ - 5^th^ (p<=0.001) visits. Despite fewer episodes of diarrhoea and fever, upper respiration infections were higher.

**Conclusion:** The comprehensive NACS demonstrated improved mother-infant nutritional and other health outcomes suggesting the need for integrated and holistic care for better maternal, infant and child health.

## Introduction

Maternal and infant malnutrition is a significant global health concern with significant implications on the overall health and well-being of both the mothers and their infants. The Global Nutrition report of 2022 [1] indicated that 29.9% of women of reproductive age suffer from anaemia while 9.1% of all women were underweight. The prevalence of low birth weights was 14.6% among the newborns while 22%, 6.7% and 5.7% of children under 5 years were stunted, wasted and overweight respectively. In the same report, Sub-saharan Africa was noted to contribute to the highest burden of malnutrition with 32.6% of children under 5year stunted, 5.2% wasted and 4% overweight while anaemia among the women of reproductive age was 31.9%. According to the Uganda Demographic Health Survey (UDHS) of 2016 [2], the prevalence of stunting among children under 5 years was 29% while underweight and wasting was 11% and 4% respectively. Additionally, aneamia affects 32% of the women of reproductive age. These surveys and reports indicate the persistent challenge of malnutrition among the women of reproductive health and children calling for urgent need for intervention and improvement.

Maternal nutrition is vital for the health and well being of both the mother and her developing infant. Maternal interventions aimed at improving nutrition practices before pregnancy, during pregnancy and lactation have been extensively studied for their potential to enhance maternal and infant health down the line [3–7]. Well nourished and healthy mothers are more likely to give birth to health babies, experience a healthy pregnancy and are less likely to experience life-threatening complications during pregnancy [8,9].

Several studies have demonstrated the positive impact of maternal interventions on the nutrition practices and growth of infants particularly when implemented in a multi-sectoral approach. For instance, implementation of a comprehensive range of interventions such as; breastfeeding promotion, education and counselling, maternal mental health, women empowerment, family planning, water, hygiene and sanitation, agricultural interventions has shown promising results in reducing stunting rates [10]. Notable studies by Olutayo et al [11], Nadia et al [12], Bhutta et al [13] emphasize the importance of these holistic interventions in reducing stunting. However, it is important to consider the perspective put forth by USAID/Advancing Nutrition [14] which argues against using stunting as a primary indicator of success of short term or single interventions at individual. Instead, stunting should be interpreted as a reflection of the population’s well-being. This view suggests a more comprehensive assessment of interventions success focusing of multiple short and long term causal factors to malnutrition instead of looking at the immediate outcomes.

Additionally, numerous studies have shown that Nutrition Counselling and education during pregnancy significantly improve maternal-infant nutrition practices as well as the overall health and nutritional status of both mothers and infants. Dearden et al [15] demonstrated a positive effect of nutrition counselling and education on maternal meal frequency and diet diversification. These findings align with a quasi-experimental study conducted by Kaleem et al [16] which indicated that Antenatal Counselling improved the maternal dietary practices and nutritional status. Similarly, a study by Perez-Escamilla et al [17] revealed a positive effect of maternal counselling on maternal and infant health and nutrition outcomes including birth weights and prevention of pre-term births. In contrast, Ghosh-Jereth et al [18] found that the maternal dietary intake remained low and anaemia rates were high despite targeted antenatal care including counselling at every visit. The authors attributed this lack of improvement to the poor quality of counselling, a finding also echoed by Nsiah-Asamoah et al [19] in their study on nutrition counselling interactions between the health workers and caregivers. While some studies showed positive effect of maternal nutrition counselling on the maternal-infant health and nutrition outcomes, contrasting findings indicate that the quality of the counselling and how it is delivered can have negative impact on these outcomes. More research is needed to bridge this gap and provide a clearer understanding on the effect of delivery of a comprehensive package including counselling on the mother-infant health and nutrition outcomes.

Therefore, providing high-quality health services, including preventive health services, antenatal, maternity and postnatal services as well as early diagnosis and treatment of medical conditions such as anaemia is crucial for improving women’s health. The World Health Organisation (WHO) and the Ministry of Health, Uganda recommends a comprehensive package of nutrition interventions to pregnant women for a positive outcome, including counselling on healthy eating and physical activity, guidance on infant and young child feeding, nutrition education on energy and protein intake, and daily iron and folic acid supplements. The package also includes energy and protein dietary supplements and high-protein supplements for the undernourished populations [20,21].

Even before the release of the WHO guidelines in 2020, the Ministry of Health in Uganda had been implementing NACS initiative, aiming to integrate nutrition into the health system and consequently improving the health and nutrition practices and outcomes of the beneficiaries. The NACS intervention package was tailored to the specific nutrition needs of the clients and was in line with WHO’s recommendations on maternal nutrition care. Support was provided to mothers and their children, covering aspects such as optimal maternal nutrition, diversified diets, iron/folic acid supplementation, iodated salt consumption, deworming, malaria prevention, and provision of antenatal care package and encouragement to attend all the 8 visits. Breastfeeding education emphasized early initiation, exclusive breastfeeding for 6 months, and extended breastfeeding. Mothers of older infants received guidance on complementary feeding practices. Caregivers of sick children were advised on continued breastfeeding. The community system offered ongoing health and nutrition care, including livelihood and economic support to improve the health and nutrition outcomes. The malnourished mothers received therapeutic foods [22].

While existing literature has assessed the impact of vertical maternal interventions on the health and nutritional status of mothers and infants, there is limited body of research on the effect of broad integrated interventions, such as NACS on the health and nutrition outcomes of beneficiaries. This study therefore, sought to assess the effect of comprehensive NACS package on the health and nutrition practices and status of mothers and their infants in Tororo and Butaleja districts in Eastern Uganda. We tested the hypotheses that there was no difference in the maternal-infant health, nutrition practices and outcomes between the facilities which integrated comprehensive NACS, verses those with routine NACS. The findings of this study contribute to the growing body of evidence on the effectiveness of broad integrated interventions on the health and nutrition outcomes of the beneficiaries and provide insights and recommendations for scaling up the NACS approach.

### Pathways on the effect of NACS integration in the health system on maternal and infant health, nutrition practices, and outcomes

Integration of comprehensive NACS into the health system will result into an integrated nutrition service delivery system which aims to foster a productive interaction between the service providers and mothers. Based on the health belief model, service providers were expected be motivated to impart knowledge and skills to mothers, enabling them take charge of their own health and nutrition. Based on the health belief model, these empowered mothers would then embrace optimal nutrition practices resulting in enhanced health and nutrition well-being. Consequently, this positive change would improve the health and nutrition outcomes of their infants, as illustrated in **S1 Fig**

## Material and Methods

### Study Design

The study used a comparative non-equivalent quasi-experimental design with two groups; comprehensive NACS integration compared to routine NACS integration.

### Study Setting and population

The study involved pregnant and lactating mothers, along with their respective infants. The two hospitals selected for the study were Tororo Hospital as the comprehensive NACS and Busolwe Hospital as the routine NACS. The hospitals were similar by level of facility, ownership, funding, staffing norms, services provided and client load. Pregnant mothers in various trimesters were enrolled and their health and nutrition status monitored at the antenatal visits and post-delivery. Only women accessing antenatal care and residing in Tororo and Butaleja were included. During post-delivery, infants were monitored for their feeding practices, health and nutrition status till 12 months.

Mothers and infants were followed through the scheduled visits at their respective health service points, which included, antenatal, labour suite/maternity, postnatal, children wards, young child and ART clinics.

### Comprehensive NACS versus routine service delivery

The comprehensive NACS package targeted both the health workers and mothers with their infants. The support to the health workers included: five-day training in NACS and Health Management Information System (HMIS) for nutrition in-service courses for staff at key health contact points, such as antenatal clinics, maternity, postnatal clinics, young child and HIV clinics; provision of anthropometric equipment, policy guidelines, job aides, information, education and communication materials for mothers; mentorships/support supervision of health staff to ensure quality service delivery; monitoring and reporting of nutrition interventions; employing quality improvement support to address gaps in NACS implementation; linking study subjects to community support structures for continuous nutrition care and support; and collaborating with key stakeholders and the district health management team to establish supervisory and support mechanisms for the intervention.

To the mothers and their infants, the package included: nutrition assessment and categorization of the nutritional status; health and nutrition education on a diversified diet, recommended antenatal clinic visits, iron/folic acid supplementation, water hygiene, and sanitation; maternal-infant nutrition counselling; provision of therapeutic feeds to identified malnourished cases; active follow-up of mother-baby pairs to ensure they receive the necessary nutrition services.

### Routine service delivery

In the routine NACS setting, some elements of NACS were integrated into the regular health care services provided such as growth monitoring and promotion for children, iron/folic acid supplementation. To ensure comparability, staff at both study settings were trained in NACS and HMIS for nutrition. They were provided with information, education and communication materials to enhance their capacity in nutrition education and counselling. Subsequently, the staff carried on with their services as per usual. The nutrition counselling placed a strong emphasis on promoting the consumption of locally available foods for the management of malnutrition. [23]

We determined the level of exposure to comprehensive verses routine NACS by closely supervising the data collection process and enhancing documentation of both the services rendered and the frequency with which the respondents accessed these services.

### Sampling

The study employed purposive sampling approach, enrolling subjects who had given their consent on a continuous basis until the desired sample size was attained. In both study settings, the antenatal care clinic served as the entry point and the ANC register as the sampling frame. The enrolment of the study participants took 8 months from starting from 23^rd^ October 2018 to 25^th^ May, 2019. The mothers were followed up till they gave birth, and the mother-infant pairs followed up for 12 months. Data was collected from 23^rd^ October, 2018 to 25^th^ July 2021.

### Sample size calculation

The sample size was calculated using the formular by V. Kasiulevicius et al [24] based on infant underweight as an outcome variable.

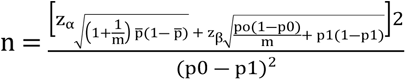

Where 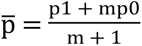

**Where,**

P_0_ was the probability of underweight infants in the control group – 0.113. P_1_was the probability of underweight infants in the intervention group – 0.07

P_0_ was based on the Uganda Demographic Health Survey 2011 burden of malnutrition in the eastern region while P1 was an estimated reduction in underweight with the intervention.

If α (alpha) = 0.05 then z_α_ = 1.96

If β (beta) = 0.80, then z_β_ = 0.845

m was the number of control subjects per experimental subject = 2

p̄ = 0.0987

n = 652 with the inclusion of 20% loss to follow up of mother-baby pairs. A sample size of 652 (217 in the intervention group and 435 in the control group) was estimated to detect a 4.3% reduction in the underweight infants at 80% power and 5% level of significance.

### Data collection

We pre-tested the data collection tools among the mothers and feedback was used to finalize the tools. The tool was designed in excel to facilitate the tracking of the mother-baby variables for their scheduled visits.

Our research assistants underwent training on the data collection tools and data capture methods at a minimum of four points: recruitment/baseline, antenatal clinic visits, delivery and postnatal care clinic, and immunization scheduled visits. We encouraged mothers to deliver at the health facility where they were provided with a package of both routine and comprehensive package of services. To ensure data quality, we conducted regular supervision and spot checks.

At baseline/recruitment, we collected data on various aspects, including socio-economic and demographic characteristics, maternal health and nutrition practices, and maternal nutritional status. Throughout each antenatal visits, we monitored the mothers’ anthropometric data, health status and nutrition practices. Following delivery, we collected data on infant’s anthropometric measurement such as birth weight, length, head circumference as well as details about their feeding practices. Subsequently, we continued to track the infants’ anthropometric data, health and nutrition practices and status during the scheduled immunization visits.

Anthropometric data and feeding practices for both the mother and her infant were collected using standard procedures. Mother’s weight was taken to the nearest 0.1gm using a digital Uniscale. The infant weight was measured to nearest 0.1 gm using the neonatal weighing scales at birth and thereafter a digital uniscale. Infant length was measured to nearest 0.1cm using an infantometer at birth and a height board for the subsequent visits.

We measured the head circumference and Mid Upper Arm Circumference (MUAC) of infants using specialized tapes, with measurements recorded to the nearest 0.1cm. MUAC was measured for infants above 6 months and mothers. Additionally, we conducted health assessment for mothers, including evaluation for illnesses such as headaches, depression, diarrhea, fever and cough. For infants, we assessed episodes of diarrhea, fever and Upper Respiratory infections.

In total, there were 15 scheduled appointments from the time of mother’s enrolment until the baby made 12 months of age. Mothers were encouraged to continue attending health facilities for continuous health care as well as participating in the informative health and nutrition education sessions.

### Data management

We used excel for data capture, STATA version 15 for data, cleaning and performing bivariate tests on all confounding background variables for both study settings. The variables included; weeks of gestation, age of the mothers, mothers’ education, marital status, mothers’ occupation, mothers’ income, spouses’ income, spouses’ education level, previous ANC visits, distance to the health facility, type of transport used, total numbers of children, number of children alive, number of family members, number of children under 5 years, fuel for cooking, water source and faecal matter disposal. We cleaned data by synchronising the variable codes for the two data sets, checked for missing data, and excluded the variables with insignificant data.

We characterised variables as continuous, binary, categorical and generated new variables. We checked the data set for normal distribution for the continuous variables. Descriptive analysis was conducted to compare mothers’ background characteristics in the 2 study arms. Continuous variables were compared using a 2 sample t-test while the categorical variables were compared using the chi square test.

### Data analysis

Because these groups were not randomly assigned and this being a quasi non-equivalent experimental study, we conducted propensity score matching to minimise potential imbalance and also create reasonable comparable groups, before assessment of the effectiveness of the NACS intervention.

By creating more comparable intervention and control groups, propensity score matching can result into a more precise estimates of intervention effects and reducing confounders. On the other hand, matching reduces the sample size, because not all individuals may find suitable matches resulting in a loss of statistical power and precision [25–27].

The propensity score matching process involved; defining the intervention (comprehensive NACS) verses control (routine NACS) groups, identification of the variables before administration of the intervention, estimating the propensity scores, checking the initial balance of the variables for both groups using mean differences, using the nearest-neighbor matching method to pair individuals in intervention and control group based on their propensity scores, assessing the quality of the matches, and thereafter estimating the effect of comprehensive NACS on maternal-infant practices, health and nutritional status [27].

Using the R software version 4.1.1, we estimated the NACS effect on maternal-infant nutrition practices, health and nutritional status by comparing various methods such as nearest neighbour, null and full matching methods. The nearest neighbour matching using the logistic regression propensity score model provided the best balance compared to other matching methods such as; full matching using a probit regression propensity score, nearest neighbour matching using a probit regression propensity score, null probit, full matching using a logistic regression propensity score as determined by the lower standardised mean difference statistics. The enrolment and data analysis flow chart is illustrated in **S2 Fig.**

### Ethical approval

This study was approved by: the Higher Degrees, Research, and Ethics Committee – Institutional Review Board at Makerere University School of Public Health (MaKSPH HDREC 24/01/2017), the Uganda National Council of Science and Technology (SS 4251), and the Office of the President in Uganda (ADM/194/212/01). An official letter from the Ministry of Health was written to the Tororo District Health Officer to seek for permission to conduct the study. The Principal Investigator informed both the District Health Officer and the District Resident Commissioner about the study plan before its execution. Participating mothers were asked to sign informed consent form. The mothers who were unable to read and write provided their informed consent using their thumbprint.

## Results

A total of 784 mothers were enrolled in the study; 423 from the Routine NACS setting and 361 from the Comprehensive NACS setting. One hundred (118) mothers were excluded from the analysis due to missing data while 666 mothers were considered in the final analysis, majority from the routine (412) compared to comprehensive (254) NACS groups.

The mothers’ characteristics at enrollment were compared in the two study arms and the results are shown in the Tables 1 and 2. The findings indicated no significant difference between the mother’s age (p= 0.466), prior antenatal visits for this pregnancy (p=0.316), number of family members (p=0.007) between the two groups at enrollment. However, there was a significant difference in the weeks of gestation (p=0.023), mothers’ income (p=0.000), spouses’ income (p=0.000), number of children (p=0.000), number of children alive (p=0.000), number of children < 5years (p=0.001), distance to health facility (p<0.001), mothers’ education (p<0.001), marital status (p<0.001), mothers’ occupation (p<0.001), spouses’ education (p<0.001), type of transport (p<0.001), cooking method (p<0.001), water source (p<0.001), and fecal matter disposal (p<0.001).

**Table 1.** Mothers’ characteristics at enrolment in the routine verses comprehensive study arms for continuous variables.

| Variable name<br>(Comprehensive NACS=254, Routine NACS = 412) | t-test | p-value |
| --- | --- | --- |
| Mothers' age | -0.730 | 0.466 |
| Weeks of gestation | 2.277 | 0.023 |
| Mothers' income | -4.682 | 0.000 |
| Spouse income | -4.997 | 0.000 |
| Prio antennal visits for this pregnancy | 1.003 | 0.316 |
| Number of children | 11.400 | 0.000 |
| Number of children alive | 5.725 | 0.000 |
| Number of family members | 2.669 | 0.007 |
| Number of children less than 5 years | 6.842 | 0.000 |

**Table 2.** Mothers’ characteristics at enrolment in the routine verses comprehensive NACS study arms for categorical variables before propensity score matching.

| Variable name | Routine NACS<br>N=412 | Comprehensive<br>NACS<br>N=254 | Chi-square Test<br>(p-Value) |
| --- | --- | --- | --- |
| **Distance to health facility** |  |  |  |
| <5 Km | 96.1% | 60.6% | <0.001 |
| >5 km | 3.9% | 39.4% |  |
| **Mothers Education** |  |  |  |
| No Education | 5.8% | 1.2% | <0.001 |
| Primary | 62.9% | 44.9% |  |
| Secondary | 26.9% | 40.6% |  |
| Higher | 4.4% | 13.4% |  |
| **Marital Status** |  |  |  |
| Married | 94.4% | 99.6% | <0.001 |
| Not married | 5.6% | 0.4% |  |
| **Mothers occupation** |  |  |  |
| Formal | 6.8% | 18.1% | <0.001 |
| Informal | 93.2% | 81.9% |  |
| **Spouse's education** |  |  |  |
| No Education | 4.2% | 1.2% | <0.001 |
| Primary | 49.3% | 25.6% |  |
| Secondary | 38.7% | 55.1% |  |
| Higher | 7.8% | 18.1% |  |
| **Type of transport** |  |  |  |
| Motorised | 50.7% | 92.1% | <0.001 |
| Walking | 49.3% | 7.9% |  |
| **Cooking method** |  |  |  |
| Firewood | 79.9% | 52.8% | <0.001 |
| Charcoal | 19.9% | 46.1% |  |
| Gas | 0.2% | 1.2% |  |
| **Water source** |  |  |  |
| Well | 0.2% | 16.1% | <0.001 |
| Borehole | 96.8% | 44.1% |  |
| Tap water | 2.9% | 39.8% |  |
| **Faecal matter Disposal** |  |  |  |
| Latrine | 99.3% | 89.0% | <0.001 |
| Toilet | 0.7% | 11.0% |  |

Propensity score matching was used to create comparability between the study groups. The Null model was used to check the initial imbalance in the two groups that the matching methods eliminated step wise. Table 3 shows severe imbalances as reflected by the standard mean differences computed by the R software. All values close to zero in the standard mean differences reflected better matches while those away from zero reflected severe imbalances. The variable ‘number of children’ had the highest difference (-4.7263) indicating severe imbalance while the variable ‘nutrition status by MUAC’ had the lowest difference (0.0019) indicating that mothers in the both groups had comparable nutritional status, which conclusions resonate with exiting literature [28].

**Table 3:** Propensity score matching null model for checking initial imbalance between the comprehensive and routine NACS study arms.

| Variables | Means Treated | Means Control | Mean difference |
| --- | --- | --- | --- |
| **Distance** | 0.8513 | 0.1053 | 3.1896 |
| **Mother age** |  |  |  |
| 15-24 | 0.4762 | 0.5140 | -0.0758 |
| 25-49 | 0.5238 | 0.4860 | 0.0758 |
| **Mothers’ education** |  |  |  |
| 1. No Education | 0.0119 | 0.0562 | -0.4082 |
| 2. Primary | 0.4444 | 0.6433 | -0.4001 |
| 3. Secondary | 0.4087 | 0.2556 | 0.3115 |
| 4. Higher | 0.1349 | 0.0449 | 0.2634 |
| **Marital status** |  |  |  |
| 1. Married | 0.9960 | 0.9438 | 0.8305 |
| 2. Not Married | 0.0040 | 0.0562 | -0.8305 |
| **Mothers’ occupation** |  |  |  |
| 1. Formal | 0.1825 | 0.0674 | 0.2980 |
| 2. Informal | 0.8175 | 0.9326 | -0.2980 |
| **Spouse education** |  |  |  |
| 1. No Education | 0.0119 | 0.0365 | -0.2269 |
| 2. Primary | 0.2579 | 0.5056 | -0.5661 |
| 3. Secondary | 0.5476 | 0.3820 | 0.3327 |
| 4. Higher | 0.1825 | 0.0758 | 0.2762 |
| **Prior ANC visits** | 2.0397 | 2.1011 | -0.0626 |
| **Distance to hospital** |  |  |  |
| 1. Less than 5km | 0.6111 | 0.9635 | -0.7228 |
| 2. ≥ 5km | 0.3889 | 0.0365 | 0.7228 |
| **Type of transport** |  |  |  |
| 1. Walking | 0.0794 | 0.5028 | -1.5665 |
| 2. Motorized | 0.9206 | 0.4972 | 1.5665 |
| **No. of children** | 1.0198 | 2.6236 | -4.7263 |
| **No. of children alive** | 1.5437 | 2.4129 | -0.5786 |
| **No of chn < 5years** | 0.7540 | 1.2809 | -0.6142 |
| **Type of fuel used** |  |  |  |
| 1. Firewood | 0.5278 | 0.7978 | -0.5408 |
| 2. Charcoal | 0.4603 | 0.2022 | 0.5178 |
| 3. Gas | 0.0119 | 0.0000 | 0.1098 |
| **Water source** |  |  |  |
| 1. Well | 0.1627 | 0.0028 | 0.4332 |
| 2. Borehole | 0.4405 | 0.9691 | -1.0648 |
| 3. Tap Water | 0.3968 | 0.0281 | 0.7537 |
| **Faecal matter disposal** |  |  |  |
| 1. Latrine | 0.8889 | 0.9916 | -0.3267 |
| 2. Toilet | 0.1111 | 0.0084 | 0.3267 |
| **Mother wtg (kg) 1<sup>st</sup> visit** | 64.2496 | 60.7542 | 0.3124 |
| **Nutritional status by MUAC** |  |  |  |
| 1. Normal (Green) | 0.9722 | 0.9719 | 0.0019 |
| 2. Malnourished (Yellow/Red) | 0.0278 | 0.0281 | 0.0019 |
| **No. of meals** | 3.0992 | 2.8792 | 0.2940 |
| **Iron folic acid supplement** |  |  |  |
| No | 0.0238 | 0.0028 | 0.1377 |
| Yes | 0.9762 | 0.9972 | -0.1377 |
| **History of headache** |  |  |  |
| No | 1.0000 | 0.8034 | 0.6465 |
| Yes | 0.0000 | 0.1966 | -0.6465 |
| **History of depression** |  |  |  |
| No | 1.0000 | 0.9803 | 0.1851 |
| Yes | 0.0000 | 0.0197 | -0.1851 |
| **History of diarrhoea** |  |  |  |
| No | 1.0000 | 0.9888 | 0.1393 |
| Yes | 0.0000 | 0.0112 | 0.1393 |
| **History of fever** |  |  |  |
| No | 0.9960 | 0.9298 | 1.0539 |
| Yes | 0.0040 | 0.0702 | -1.0539 |
| **History of cough** |  |  |  |
| No | 1.0000 | 0.9522 | 0.2927 |
| Yes | 0.0000 | 0.0478 | -0.2927 |

### Effect NACS integration on the mothers-infant health, nutrition practices and status

The study assessed the effect of NACS integration on maternal-infant nutrition practices as well as its effects on health and nutritional status. This assessment employed the nearest neighbour matching method along with a logistic regression propensity score model and the results are shown in Table 4.

**Table 4.** The effect of NACS integration on the mother-infant nutrition practices, health and nutrition status using nearest neighbour matching logistic regression propensity score model.

| Variable name | Contrast |  | Estimate | SE | P value | CI |
| --- | --- | --- | --- | --- | --- | --- |
|  | 1-(Comp<br>NACS) | 0-(Routine<br>NACS) |  |  |  |  |
| **Mother variables** | N=252 | N=252 |  |  |  |  |
| **Meal frequency at 2<sup>nd</sup> visit** | 1 | 0 | 0.008 | 0.070 | 0.911 | -0.13, -0.146 |
| **Diet Diversity Score** |  |  |  |  |  |  |
| 2<sup>nd</sup> visit | 1 | 0 | -3.110 | 0.263 | <0.001 | -3.630, - 2.600 |
| 3<sup>rd</sup> visit | 1 | 0 | -2.980 | 0.213 | <0.001 | -3.400, -2.560 |
| 4<sup>th</sup> visit | 1 | 0 | -2.690 | 0.149 | <0.001 | 2.990, -2.400 |
| **Iron/ folic acid supplementation** |  |  |  |  |  |  |
| 2<sup>nd</sup> visit | 1 | 0 | -0.029 | 0.010 | 0.006 | -0.049, -0.008 |
| 3<sup>rd</sup> visit | 1 | 0 | -0.024 | 0.010 | 0.012 | -0.043 , -0.005 |
| 4<sup>th</sup> visit | 1 | 0 | -0.035 | 0.015 | 0.017 | -0.064 , -0.006 |
| **Weight at** |  |  |  |  |  |  |
| 2<sup>nd</sup> visit | 1 | 0 | 1.040 | 0.272 | <0.001 | 0.507 -1.570 |
| 3<sup>rd</sup> visit | 1 | 0 | 2.690 | 0.422 | <0.001 | 1.860 – 3.520 |
| 4<sup>th</sup> visit | 1 | 0 | 5.860 | 1.990 | 0.003 | 1.950 – 9.760 |
| **Nut.status by (MUAC)** |  |  |  |  |  |  |
| 2<sup>nd</sup> visit | 1 | 0 | 0.038 | 0.018 | 0.032 | 0.003- 0.072 |
| 3<sup>rd</sup> visit | 1 | 0 | 0.026 | 0.009 | 0.047 | 0.008 -0.044 |
| 4<sup>th</sup> visit | 1 | 0 | 0.025 | 0.015 | 0.091 | 0.004- 0.054 |
| **History of headache** |  |  |  |  |  |  |
| 2<sup>nd</sup> visit | 1 | 0 | 0.020 | 0.009 | 0.020 | 0.003 - 0.037 |
| 3<sup>rd</sup> visit | 1 | 0 | 0.044 | 0.013 | <0.001 | 0.020- 0.069 |
| 4<sup>th</sup> visit | 1 | 0 | 0.078 | 0.022 | <0.001 | 0.034 - 0.121 |
| **History of depression** |  |  |  |  |  |  |
| 2<sup>nd</sup> visit | 1 | 0 | 0.013 | 0.007 | 0.070 | -0.001, -0.026 |
| 3<sup>rd</sup> Visit | 1 | 0 | 0.008 | 0.005 | 0.160 | -0.003, -0.018 |
| **Diarrhoea at 3<sup>rd</sup> visit** | 1 | 0 | 0.018 | 0.008 | 0.030 | 0.002 -0.034 |
| **History of fever** |  |  |  |  |  |  |
| 2<sup>nd</sup> visit | 1 | 0 | 0.020 | 0.009 | 0.019 | 0.003 - 0.037 |
| 3<sup>rd</sup> visit | 1 | 0 | 0.049 | 0.014 | <0.001 | 0.023 - 0.076 |
| 4<sup>th</sup> visit | 1 | 0 | 0.030 | 0.013 | 0.021 | 0.005 - 0.055 |
| **Infant variables** |  |  |  |  |  |  |
| **Infant birth weight** | 1 | 0 | 0.191 | 0.100 | 0.056 | -0.005, -0.387 |
| **Infant weight at** |  |  |  |  |  |  |
| 3<sup>rd</sup> visit | 1 | 0 | 1.050 | 0.111 | <0.001 | 0.835 - 1.270 |
| 4<sup>th</sup> Visit | 1 | 0 | 1.130 | 0.106 | <0.001 | 0.923 - 1.340 |
| 5<sup>th</sup> visit | 1 | 0 | 1.930 | 0.107 | <0.001 | 1.720 - 2.140 |
| **Infant head cirm** |  |  |  |  |  |  |
| 3<sup>rd</sup> visit | 1 | 0 | 3.900 | 0.248 | <0.001 | 3.410 - 4.380 |
| 4<sup>th</sup> visit | 1 | 0 | 3.900 | 0.248 | <0.001 | 3.410 - 4.380 |
| 5<sup>th</sup> visit | 1 | 0 | 0.020 | 0.009 | 0.019 | 0.003 -0.037 |
| **Wt for Age Z-scores** |  |  |  |  |  |  |
| 3<sup>rd</sup> visit | 1 | 0 | 1.800 | 0.233 | <0.001 | 1.350-2.260 |
| 4<sup>th</sup> visit | 1 | 0 | 1.700 | 0.208 | <0.001 | 1.290 -2.110 |
| 5<sup>th</sup> visit | 1 | 0 | 2.730 | 0.202 | <0.001 | 2.330 – 3.130 |
| 6<sup>th</sup> visit | 1 | 0 | 0.265 | 0.159 | 0.096 | -0.047, -0.577 |
| **Length for Age Z-scores** |  |  |  |  |  |  |
| 4<sup>th</sup> visit | 1 | 0 | 0.634 | 0.246 | 0.010 | 0.151 – 1.120 |
| 5<sup>th</sup> Visit | 1 | 0 | 0.761 | 0.222 | <0.001 | 0.326 – 1.200 |
| 6<sup>th</sup> visit | 1 | 0 | 3.990 | 0.281 | <0.001 | 3.440 – 4.540 |
| 7<sup>th</sup> visit | 1 | 0 | 4.630 | 1.140 | <0.001 | 2.470 – 6.780 |
| **Wt. for Length Z-scores at** |  |  |  |  |  |  |
| 3<sup>rd</sup> visit | 1 | 0 | 6.250 | 0.475 | <0.001 | 5.320 – 7.180 |
| 4<sup>th</sup> visit | 1 | 0 | 1.460 | 0.356 | <0.001 | 0.763 – 2.160 |
| 5<sup>th</sup> visit | 1 | 0 | 2.770 | 0.342 | <0.001 | 2.100 – 3.440 |
| **History of Upper Respiratory Infection** |  |  |  |  |  |  |
| 5<sup>th</sup> Visit | 1 | 0 | 0.043 | 0.013 | <0.001 | 0.018 - 0.069 |
| 6<sup>th</sup> visit | 1 | 0 | 0.155 | 0.023 | <0.001 | 0.110 - 0.199 |
| 7<sup>th</sup> visit | 1 | 0 | 0.294 | 0.029 | <0.001 | 0.237 - 0.350 |
| 8<sup>th</sup> visit | 1 | 0 | 0.315 | 0.029 | <0.001 | 0.258 - 0.372 |
| **History of infant diarrhoea** |  |  |  |  |  |  |
| 5<sup>th</sup> visit | 1 | 0 | -0.168 | 0.080 | 0.036 | -0.324, -0.011 |
| 6<sup>th</sup> visit | 1 | 0 | -0.146 | 0.080 | 0.067 | -0.302- 0.010 |
| 7<sup>th</sup> visit | 1 | 0 | -0.033 | 0.049 | 0.506 | -0.130- 0.064 |
| 8<sup>th</sup> visit | 1 | 0 | -0.126 | 0.067 | 0.061 | -0.130- 0.064 |
| **History of infant fever** |  |  |  |  |  |  |
| 1<sup>st</sup> visit | 1 | 0 | -0.054 | 0.030 | 0.072 | -0.113- 0.005 |
| 3<sup>rd</sup> visit | 1 | 0 | -0.005 | 0.012 | 0.694 | -0.029- 0.019 |
| 4<sup>th</sup> visit | 1 | 0 | -0.409 | 0.086 | <0.001 | -0.576, -0.241 |
| 5<sup>th</sup> Visit | 1 | 0 | -0.111 | 0.076 | 0.145 | -0.260- 0.038 |
| 6<sup>th</sup> visit | 1 | 0 | -0.023 | 0.062 | 0.712 | -0.144 - 0.099 |
| 7<sup>th</sup> visit | 1 | 0 | 0.028 | 0.050 | 0.569 | -0.070 - 0.126 |
| 8<sup>th</sup> visit | 1 | 0 | -0.027 | 0.060 | 0.657 | -0.143- 0.090 |

Mothers in both groups were similar in terms of; meal frequency (p=0.911), iron/folic acid supplementation at the 2^nd^ -4^th^ visits (β <= -0.035), maternal nutritional status by MUAC at the 2^nd^ – 4^th^ visits (β<= 0.038). Additionally there was no significant difference in maternal instances of; headache at the 2^nd^ -4^th^ visits (β<=0.078), depression at the 2^nd^ -3^rd^ visit (β<=0.013) and diarrhoea at the 2^nd^ -4^th^ (β<=0.049) visits.

Whereas mothers in the routine NACS group had a significantly higher diversity score at the 2^nd^ - 4^th^ visits (p<0.001), the comprehensive group had higher weights at the 2^nd^ - 4^th^ (p<=0.003) visits. The difference in weights increased with number of visits right from the time of mothers’ enrolment.

Compared to routine, infants born to mothers in the comprehensive group had a significantly higher; birth weights at 10% level of significance (p=0.056, CI -0.005 – 0.387), weight-for-age at the 3^rd^ -6^th^ visits (p<0.001) with 20% reduction in underweight on average per visit. Furthermore, their length-for-age was significantly higher at the 4^th^ -7^th^ visits (p<0.001). The difference widened with the increasing number of visits. Similarly, the weight-for-length of the comprehensive NACS group was significantly much higher at the 3^rd^ -5^th^ visits (p<0.001). The difference remained constant throughout the subsequent visits.

Unlike the routine group, infants in the comprehensive NACS group had significantly higher episodes of upper respiratory infections at the 5^th^ -8^th^ (p<0.001, β<=0.315). On the other hand, the routine NACS infants experienced a significantly higher episodes of; diarrhoea at the 5^th^ - 8^th^ visits (p<=0.061) and fever at the 1^st^ (p=0.072, β=-0.054) and 4^th^ visits (p<0.001, β=-0.409).

## Discussion

The study aimed to assess the effect of NACS integration on the maternal-infant nutrition practices, health and nutritional status. The findings provide insights into the potential benefits of nutrition integration on the health system on the wellbeing of the mothers and their infants. The key findings in light of existing evidence and their implication are discussed.

The study found no significant difference in meal frequency among the mothers in both study groups, suggesting similar dietary habits. Compared to the comprehensive group, mothers in the routine NACS setting had a significantly higher diversity scores on all visits. This disparity may be attributed to the close proximity to the rural settings offering more natural and diverse food choices. Maternal nutritional status by MUAC estimates exhibited no significant differences across the visits in the two study settings. In contrast to the routine group, mothers in the comprehensive displayed significantly higher weights at the 2nd-4th visits. This implied that integration of comprehensive NACS had positive progressive impact on maternal weight gain from the time of enrolment. The findings suggest potential program implication and highlight the need to consider environmental context when implementing nutrition programs. Future nutrition interventions should therefore be tailored to the specific needs and context of the target population. Furthermore, the study re-enforces, existing body of evidence indicating that maternal focused interventions particularly those with a multi-sectoral nature contribute to improved maternal diet diversity, micronutrient intake and overall nutritional status [29–31].

Additionally, there were minimal difference in iron/folic acid supplementation between the two groups at the 2^nd^ -4^th^ visits implying consistent adherence to the Ministry of Health guidance on routine iron/folic acid supplementation among pregnant mothers in both settings. However, it is worth noting that the effect of iron/folic acid supplementation on haemoglobin levels in both settings could not be assessed in both settings due to lack of equipment and supplies. Studies by Michael Habtu et al [32], Sunita Taneja et al [7], Melesse Kuma et al [33] revealed elevated haemoglobin levels among women in the intervention group, findings that our study was unable to replicate due to the constraints related to equipment and supplies.

The estimates showed no difference between the two settings for maternal episodes of headache, depression and diarrhoea across the various visits. This implies that these health concerns are common and not influenced by the study settings. These need to be addressed in both setting for the well being of mothers.

Our investigation into the nutritional status of the infants revealed that integration of comprehensive NACS increased infant birth weights, reduced instances of underweight, stunted and wasted infants. This implies that nutrition integration had a potential benefit of on the foetal and infant growth and development. Our findings concur with; Veeena et al [34] M Barker et al [8], Von Salmuth et al [10], Olutayo et al [11], Micheal Habtu etal [35] on the effectiveness of a holistic approach to improving the nutritional status of children.

Routine NACS infants experienced significantly more episodes of diarrhea and fever at the various visits than the comprehensive NACS group. The findings concur with Gonzalenz-Fernandez et al [36] in their study in which implementation of the multisectoral approach lowered the risk of diarrhoea and respiratory infections. This implies that the health facilities in the routine NACS settings did not comprehensively address these health concerns hence the need for more interventions for better health and nutrition outcomes.

One of the strengths of this study lies in its comparison of two separate groups; routine versus comprehensive and its close monitoring of the practices and outcomes of the study participants. This approach bolstered the study’s findings, providing a clear and robust insight into the effectiveness of the integrated intervention package. Moreover the study places emphasis on favourable outcomes of comprehensive NACS highlighting the potential benefits of such comprehensive interventions, which findings are also consistent with the existing literature. On the other hand, the study lacked the ability to assess the impact of iron/folic acid supplementation on haemoglobin levels due to lack of equipment and supplies, which is a limitation in understanding the complete maternal health and nutrition outcomes.

## Conclusions

The findings add to the existing body of evidence supporting improved maternal-infant health and nutrition practices and status with integrated nutrition services. While meal frequency, iron/folic acid supplementation were similar in the both groups, integration of comprehensive NACS intervention improved; maternal weights, infant birth weights, infant growth in light of weight-for-age, length-for-age and weight-for-length. This emphasises the potential benefits of integrated nutrition interventions in promoting the overall being of the mothers and their infants.

## Recommendations

Based on the above findings, the Ministry of Health should consider: investing in acquiring the necessary equipment and supplies to assess the impact of iron/folic acid supplementation on haemoglobin levels for comprehensive evaluation of the women; scaling up integration of the comprehensive NACS in the health system as it has positive effect on the maternal-infant nutrition practices, health and nutrition outcomes; investing in digitization to ease monitoring and tracking trends in the health and nutrition status of the mother-infant pairs.

Future research can focus on implementation and effectiveness of digitization in monitoring and tracking of the maternal-infant health and nutritional status in an integrated health system. Secondly, it will be important to investigate the experiences of the women and caregivers receiving these services.

## Data Availability

All relevant data are within the manuscript and its supporting information files

## Supporting information

S1 File Dataset for Tororo and Butaleja districts

S1 Fig. Pathways on the effect of NACS integration into the health system on maternal and infant health, nutrition practices, and outcomes adapted from the Chronic Care Model

S2 Fig. Enrolment and data analysis flow chart

## Acknowledgements

We extend our appreciation to the political and administrative authorities of Tororo and Butaleja districts for granting us the necessary permission to conduct this study. Our gratitude go to the health workers as well as the mothers and their infants from Tororo and Butaleja districts for the dedicated participation in advancing this research undertaking. Brian Wakoli is appreciated for the statistical support rendered during the analysis.

## Author contribution

**Conceptualization:** SN FEM GWM SNK

**Data Curation:** SN FEM SNK

**Formal analysis:** SN SNK FEM

**Funding acquisition:** SN

**Investigation:** SN

**Methodolog**y: SN SNK FEM

**Project administration:** SN

**Resources:** SN

**Software:** SN

**Supervision:** SN FEM GWM SNK

**Validation:** SN FEM GWM SNK

**Visualization:** SN FEM

**Writing-Original draft**: SN

**Writing – Review and editing:** SN FEM GWM SNK

